# Fecal microbiota transplantation increases colonic IL-25 and dampens tissue inflammation in patients with recurrent *Clostridioides difficile*

**DOI:** 10.1101/2021.07.16.21260643

**Authors:** Ning-Jiun Jan, Noah Oakland, Pankaj Kumar, Girija Ramakrishnan, Brian W. Behm, William A. Petri, Chelsea Marie

## Abstract

**Background:** *Clostridioides difficile* infection (CDI) is the most common hospital-acquired infection in the United States. Antibiotic-induced dysbiosis is the primary cause of susceptibility and fecal microbiota transplantation (FMT) has emerged as an effective therapy for recurrence. We previously demonstrated in the mouse model of CDI that antibiotic-induced dysbiosis reduced colonic expression of IL-25, and that FMT protected in part by restoring gut commensal bacteria-mediated IL-25 signaling. Here we conducted a prospective clinical trial to test the impact of FMT on immunity, specifically testing in humans if FMT induced IL-25 expression in the colon.

**Methods:** Subjects received colonic biopsies and blood sampling at the time of FMT and 60-days later. Colon biopsies were assayed for IL-25 by immunoassay, for mRNA by RNAseq, and for bacterial content by 16 S rDNA sequencing. High dimensional flow cytometry was also conducted on peripheral blood mononuclear cells pre- and post-FMT.

**Results:** All 10 subjects who received FMT had no CDI recurrences over a 2 year follow-up post FMT. FMT increased alpha diversity of the colonic microbiota and was associated with several immunologic changes. The cytokine IL-25 was increased in colonic tissue. In addition, increased expression of homeostatic genes and repression of inflammatory genes was observed in colonic mRNA transcripts. Finally, circulating Th17 cells were decreased post-FMT.

**Conclusion:** The increase in the cytokine IL-25 accompanied by decreased inflammation is consistent with FMT acting in part to protect from recurrent CDI via restoration of commensal activation of type 2 immunity.

## Introduction

*Clostridioides difficile* (*C. difficile*) is an opportunistic pathogen that can cause life-threatening diarrhea and colitis. Initial *C. difficile* infection (CDI) is typically treated with antibiotics such as vancomycin or fidaxomycin (McDonald, Gerding et al. 2018). Efficacy of standard antibiotic therapy for primary CDI is 58-78% and decreases substantially after the first recurrence of disease (Oldfield, Oldfield et al. 2014). Approximately 20-30% of patients develop recurrent CDI (rCDI) within 2 weeks of completion of therapy (Deshpande, Pasupuleti et al. 2015). Fecal microbiota transplant (FMT) is an effective treatment for rCDI: a recent meta-analysis found the overall efficacy of FMT to be 76.1% with lower efficacy for treating patients with refractory CDI as opposed to rCDI (63.9% vs 79%)(Tariq, Pardi et al. 2019).

Despite the increasing use of FMT for treatment of rCDI, the mechanisms of action of FMT are poorly defined. It is hypothesized that FMT has direct inhibitory effects on *C. difficile* via niche exclusion, nutrient competition and the production of antimicrobial peptides(Madan and Petri Jr 2012, Jenior, Leslie et al. 2017, Fletcher, Pike et al. 2021). FMT may also induce changes in the host intestinal epithelium that increase resistance to recurrence of disease via fortification of the mucus layer, and differentiation and proliferation of intestinal epithelial cells (Zheng, Liwinski et al. 2020).

One reason the mechanisms of FMT are unclear is that host immune responses can vary greatly and have complex effects on *C. difficile* infection severity, as well as the efficacy of FMT (Pantosti, Cerquetti et al. 1989, Péchiné, Janoir et al. 2005, Abt, Littmann et al. 2020, Frisbee and Petri 2020). Type-1 responses via ILC1s have been shown to be protective (Abt, Lewis et al. 2015), type-17 immune responses have been related to increased host damage, and type-2 immune responses via ILC2s have been related to tissue repair via eosinophil recruitment. (Buonomo, Cowardin et al. 2016, Frisbee, Saleh et al. 2019) It is therefore important to consider patient variability and the interplay between microbiota and host immune-response when analyzing the mechanisms behind FMT as a treatment for rCDI.

We hypothesized that FMT acts to protect from rCDI in part by restoring commensal bacterial signaling to the innate immune system via the intestinal epithelial produced cytokine IL-25. To test this hypothesis, subjects with rCDI undergoing FMT were prospectively enrolled in a clinical study and colonic biopsies and peripheral blood were collected pre-and post-FMT. The goal was to analyze the local and peripheral immune responses induced by FMT. Colonic biopsies were used to assess the transcriptional response of the intestinal epithelium to FMT by RNA-sequencing and the protein-level response using immunoassays for cytokines. Intestinal and peripheral immune cell populations were analyzed by high dimensional flow cytometry. In addition, host epithelial-associated bacteria were identified by 16 S rDNA sequencing from the colonic biopsies.

## METHODS

### Study Design

The study is registered in ClinicalTrials.gov (Identifier: NCT02797288). The goal of this study was to identify the immune mechanisms underlying successful FMT for rCDI. Specifically, we tested if FMT increased IL-25 signaling, which has been identified as initiating a protective type 2 immune response in the mouse model of CDI. Participants were recruited from patients undergoing FMT treatment for recurrent *C. difficile* infection as a part of their medical care at the University of Virginia Health System (UVAHS). All patients underwent standard vancomycin antibiotic treatment prior to FMT. Colonic biopsies and whole blood were collected immediately prior to FMT and 60 days after FMT. A total of 10 participants were recruited (Table 1). Biopsies were obtained from the sigmoid colon in subjects at the time of fecal transplantation. Follow-up biopsies were obtained from the sigmoid colon 60 days after FMT for six subjects. The primary outcome measure was colonic tissue IL-25 concentration. It was hypothesized that successful FMT would restore IL-25 and IL-4 in the colon. Biopsies taken for research purposes at each colonoscopy were analyzed to determine tissue levels of immune signaling proteins IL-25, IL-4, and IL-1, gene expression by RNA-sequencing, infiltrating and circulating immune cells using high dimensional flow-cytometry, and microbiota changes by 16s rDNA sequencing.

**Table 1.** Demographics and Baseline Clinical Data of Patients that underwent FMT treatment for recurrent *C. difficile* infection.

| Clinical Characteristics | Total Cohort<br>n = 10 | Cohort with Second Visit<br>n = 6 |
| --- | --- | --- |
| Second Visit, percent (No./Total) | 60% (6/10) | 100% (6/6) |
| Age at FMT, mean (SD), y | 71 (7.1)~ | 69.83 (8.70) |
| Women, percent (No./Total) | 90% (9/10) | 100% (6/6) |
| BMI at FMT, mean (SD)* | 31.76 (8.91)~ | 28.79 (8.68) |
| No. of CDI recurrences per patient pre FMT, mean (SD) | 3.1 (0.60) | 3.2 (0.75) |
| 2, percent (No./Total) | 10% (1/10) | 17% (1/6) |
| 3, percent (No./Total) | 70% (7/10) | 50% (3/6) |
| 4, percent (No./Total) | 20% (2/10) | 33% (2/6) |
| Treated with vancomycin prior to FMT | 100% (10/10) | 100% (6/6) |
| Developed IBS-like symptoms Post FMT | 20% (2/10) | 33.3% (2/6) |
| Recurrences post FMT | 0% (0/10) | 0% (0/6) |
| No. of CDI recurrences post FMT | 0% (0/10) | 0% (0/6) |
| Same FMT donor as another subject: |  |  |
| Yes, percent (No./Total) | 40% (4/10) | 50% (3/6) |
| No, percent (No./Total) | 60% (6/10) | 50% (3/6) |

~missing data for one patient

**Table 2.**
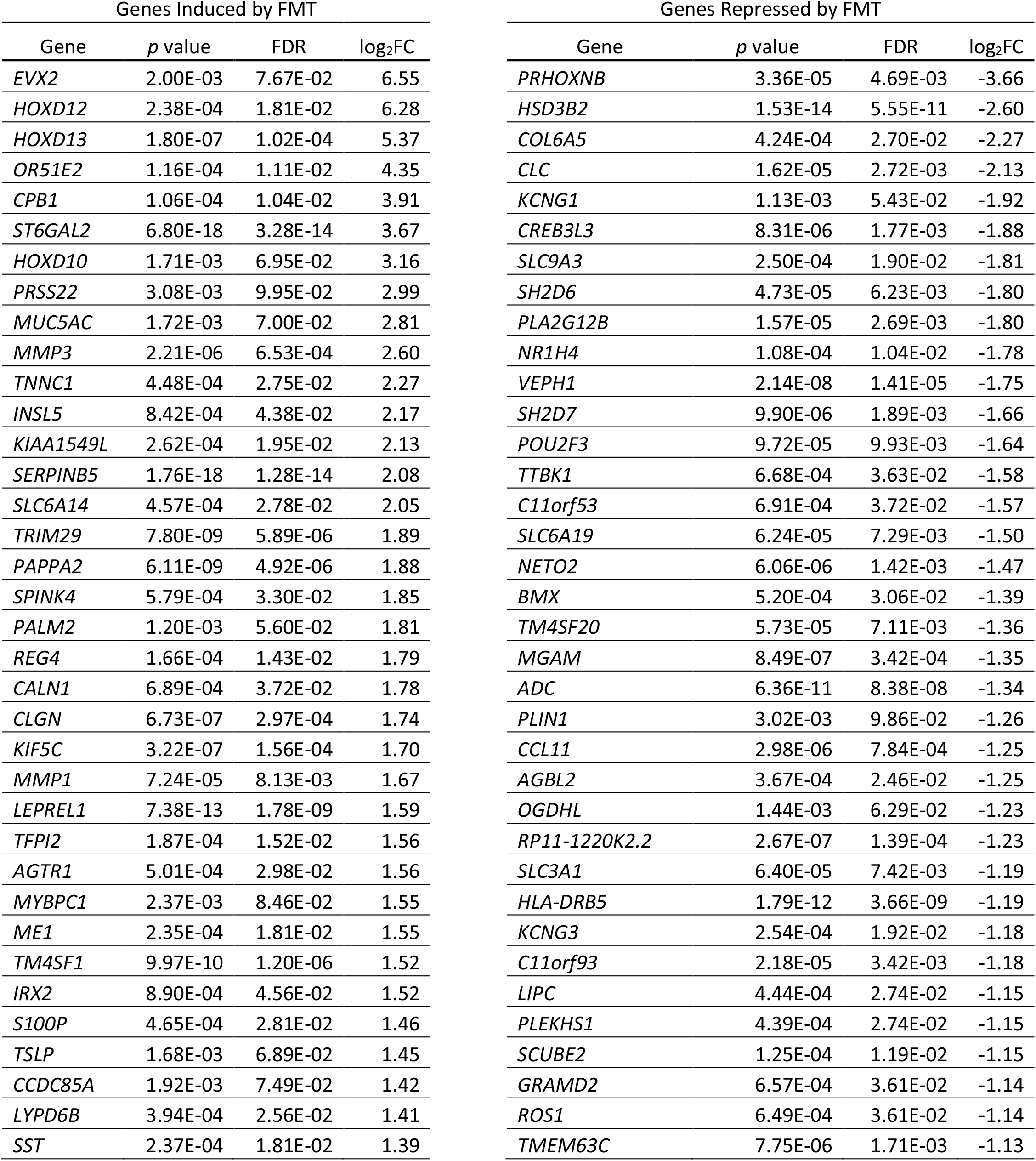
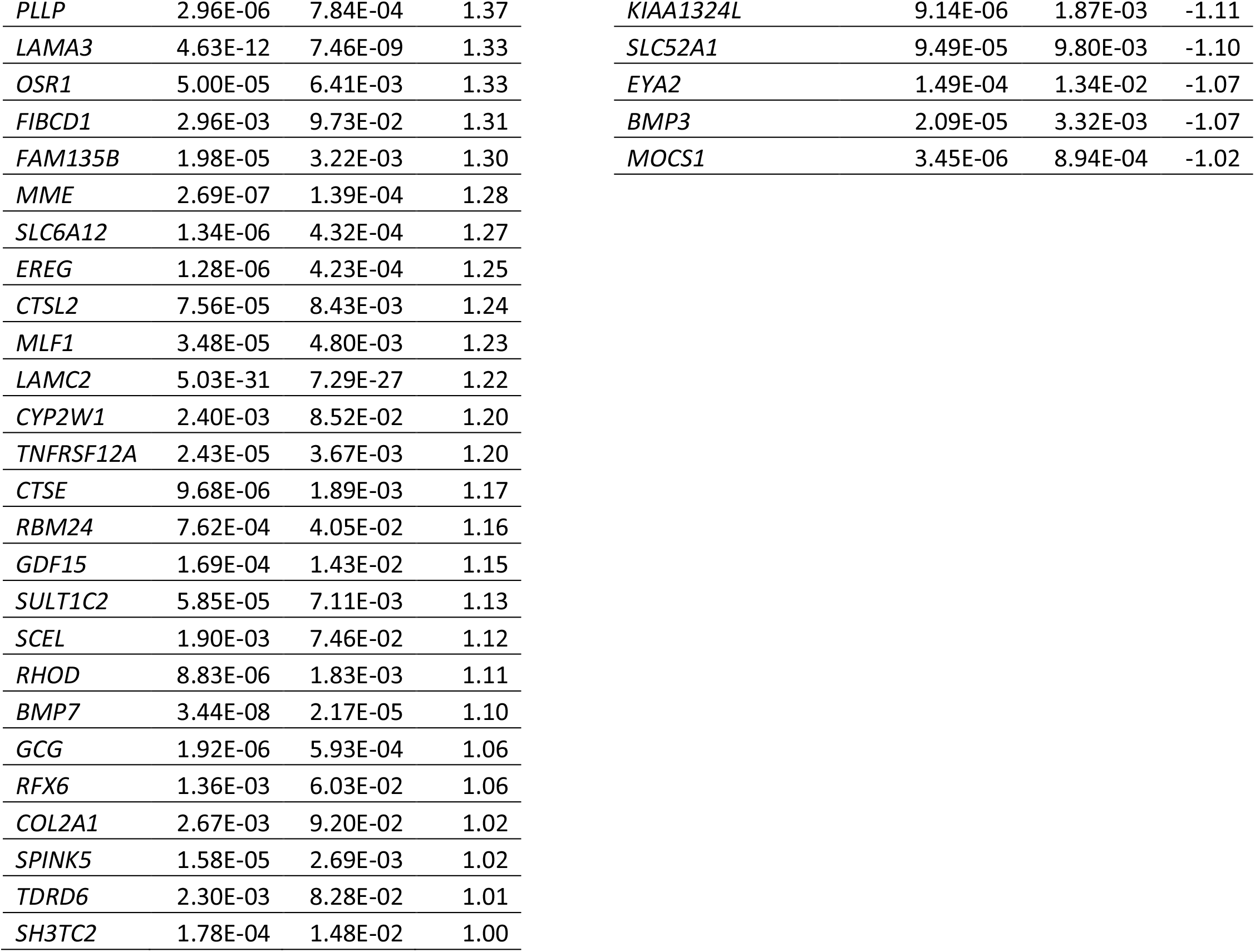
Differentially expressed genes between pre and post FMT. A Wald test was used to test for significance and adjusted for multiple comparisons using the Benjamini-Hochberg method. Genes are separated into induced (left) and repressed (right) and ordered by increasing decreasing absolute log_2_FC.

### Luminex Immunoassay

Colonic biopsies were immediately stored at -80° C, dry, until protein extraction. The biopsies were lysed in MAP lysis buffer (Millipore) with HALT proteinase inhibitors (ThermoFisher) using 5 mm steel beads (Qiagen) with a bead-beater (Tissuelyzer II, Qiagen). After centrifugation, the supernatants were transferred to -80° C until use in immunoassays. The lysates were thawed and a 47-plex magnetic bead-based Luminex immunoassay kit (Milliplex, Millipore EMD) was used to measure protein concentrations. The mean fluorescence intensity (MFI) for each protein was read using Luminex MagPix and analyzed using Milliplex Analyst 5.1 (Millipore EMD), which uses a 5-log standard fit to calculate protein concentration from MFI. For MFIs outside the range of the standards, the concentrations were extrapolated using their respective standard curves (derived from the Milliplex Analyst, back calculated using Matlab, Mathworks). Differences in protein concentrations between pre and post FMT were tested for significance (alpha = 0.05) using a linear mixed effect model, where patient ID was included as a random variable (Table S2). IL-1a, IL-1b, IL-4, and IL-25 were defined a priori in the analysis plan so the p-values were not corrected for multiple comparisons. A Benjamini-Hochberg adjustment was used for the p-values calculated for the 43 remaining proteins since they were not pre-selected.

### Messenger RNA-Seq

Colonic biopsies were obtained during colonoscopy and immediately placed in allprotect (Qiagen) and stored at -80 C until nucleic acid extraction (Qiagen AllPrep). A total amount of 1 μg RNA per sample was used as input material for the RNA sample preparations. Sequencing libraries were generated using NEBNext Ultra RNALibrary Prep Kit for Illumina (NEB, USA) following manufacturer’s recommendations. Briefly, mRNA was purified from total RNA using poly-T oligo-attached magnetic beads. Fragmentation was carried out using divalent cations under elevated temperature in NEBNext First Strand Synthesis Reaction Buffer (5X). First strand cDNA was synthesized using random hexamer primer and M-MuLV Reverse Transcriptase (RNase H-). Second strand cDNA synthesis was subsequently performed using DNA Polymerase I and RNase H. Remaining overhangs were converted into blunt ends via exonuclease/polymerase activities. After adenylation of 3’ ends of DNA fragments, NEBNext Adaptor with hairpin loop structure were ligated to prepare for hybridization. In order to select cDNA fragments of preferentially 150∼200 bp in length, the library fragments were purified with AMPure XP system (Beckman Coulter, Beverly, USA). Then 3 μl USER Enzyme (NEB, USA) was used with size-selected, adaptor ligated cDNA at 37 °C for 15 min followed by 5 min at 95 °C before PCR. Then PCR was performed with Phusion High-Fidelity DNA polymerase, Universal PCR primers and Index (X) Primer using the NEBNext Multiplex Oligos for Illumina. PCR products were purified (AMPure XP system) and library quality was assessed on the Agilent Bioanalyzer 2100 system. Clustering of index-coded samples was performed on a cBot Cluster Generation System using PE Cluster Kit cBot-HS (Illumina) according to the manufacturer’s instructions. After cluster generation, the library preparations were sequenced on a MiSeq (Illumina) and 150 bp paired-end reads were generated.

### mRNA-Seq Analysis

A total of 13 samples were processed, 1 from pre FMT and 1 from post FMT for each of 6 patients except for patient 3 which had 2 samples from pre FMT as a control for batch effects. Raw reads were processed through fastp (Chen, Zhou et al. 2018) to remove adapter, poly-N sequences, and reads with low quality. Q20, Q30 and GC content of the clean data were calculated, and only high-quality reads were preserved. Paired-end clean reads were aligned to the reference genome Homo Sapiens (GRCh37/hg19) using the Spliced Transcripts Alignment to a Reference (STAR) software (Dobin, Davis et al. 2013). FeatureCounts was used to quantify reads mapped to each gene (Liao, Smyth et al. 2014). Read counts were processed using the bioconductor package DESeq2 v1.30.1 in R (version 4.0.5) and normalized using the DESeq algorithm. Principal component analysis (PCA), hierarchical clustering, and density maps(Kolde and Kolde 2015) assessed overall similarity between samples and was used to determine the importance and influence of various factors, including the effects of FMT, patient, and persistent IBS symptoms following FMT. From the PCA, top positive and negative loadings for components 1 and 2 were reported. For the density maps, a heatmap was constructed from the top 30 most variable genes. For each gene, the standard deviation of counts across all samples was calculated as a measure of variability. Differentially expressed genes were calculated using the Wald test in DESeq2 (Love, Anders et al. 2014). We used two models, a grouped model and a patient-specific model. The grouped model contrasted between pre and post FMT while controlling for patient effects by adding patient ID as a separate variable to the model (design = ∼Patient+Visit) (n=13). The patient-specific model contrasted pre and post FMT for each paired patient (6 total) by using a term combining patient ID and whether the sample was pre or post FMT (design = ∼Patient_Visit). Genes with log_2_fold change differences (FC) ≥ 1 and using the Benjamini–Hochberg false discovery rate correction (FDR, 0.1) were considered significant.

### Gene Set Enrichment Analysis

Gene set enrichment analysis was performed on a list of all genes and their respective log2FCs in the grouped model. The package fgsea v3.13. (Korotkevich, Sukhov et al. 2021) in R was used to analyze the fold changes of the genes to identify enriched gene pathways and functions using KEGG pathways. P values are calculated using an enrichment score statistic (Korotkevich, Sukhov et al. 2019). Enrichment maps were constructed to determine functional modules using the emapplot function within the enrichplot v1.10.1 package () in R. The resulting enriched pathways and their corresponding gene lists were then selectively narrowed for each patient using the patient-specific model to measure the extent of enrichment for a pathway for each patient. As a measure of overall FC, the log FCs for the differentially expressed genes were summed for each enriched gene pathway of interest for each patient with paired RNA-seq data between pre and post FMT. This sum represented the extent of enrichment for a given pathway for that patient.

### Flow Cytometry of LPMCs and PBMCs

Lamina propria mononuclear cells (LPMCs) were isolated from colonic biopsies as follows. Colonic biopsies were submerged in ice-cold HBSS without calcium and magnesium and immediately transported to the laboratory for cell disassociation. The biopsies were digested at room temperature for 20 minutes (Accumax, eBiosciences), filtered (70 µm), to isolate LPMCs. For PBMC isolation, whole blood was centrifuged, and the plasma removed. The PBMCs were isolated by adding Ficoll (Cytiva) and then centrifuged in SepMate tubes (Stemcell Technologies). Isolated LPMCs and PBMCs were cryopreserved in liquid nitrogen until staining.

Isolated LPMCs and PBMCs were thawed and stained with fluorochrome-conjugated antibodies (Table S1). Appropriate controls (unstained, single stained, and fluorescence minus one (FMO) stained PBMCs as controls. In addition, each sample also underwent a fixed viability stain (Zombie NIR, Biolegend). Cells were first stained with surface stains, then permeabilized (Foxp3 Transcription Factor Staining Buffer Set, Invitrogen) for intracellular staining. Samples were analyzed with the 5 Laser Cytek Aurora Borealis flow cytometer. All cells were collected for LPMC samples and 100,000 cells for PBMC samples. These fluorescence reads were then analyzed using FlowJo V.10.7.2. to phenotype immune cell subsets via tSNE clustering and traditional gating (Figure S2). Spectral deconvolution and gating were based on single stained and fluorescence minus one (FMO) stained PBMC control samples.

For each tSNE cluster or immune cell phenotype of any given sample, the raw counts and percentage of leukocyte population (viable CD45+ cells) was calculated. Differences in these raw counts and percentages between pre and post FMT were quantified using a linear mixed effect model, where patient ID was included as a random variable.

### 16 S rDNA Sequencing and analysis of epithelial-associated microbiota

Bacterial DNA from colonic biopsies was amplified using V4 specific primers and indexed using Nextera XT Index Kit, which was then sequenced by Illumina via MiSeq sequencing (Kozich, Westcott et al. 2013). The amplification was insufficient from one post FMT sample, narrowing the number of samples analyzed (10 pre FMT and 5 post FMT). A mock sample (ZymoBIOMICS) was included as a control. The resulting library was pre-processed through the DADA2 package (Callahan, McMurdie et al. 2016) in R to form a library of amplicon sequence variants (ASVs). Taxonomy was assigned with Silva (Quast, Pruesse et al. 2012) release 138 databases. Forward reads were truncated at 240bp while reverse reads were truncated at 160bp. A paired-end library was used with expected amplicon size of around 250bp. This length was confirmed after merging forward and reverse reads in the DADA2 pipeline.

Alpha diversity measures Shannon and Simpson diversity values were calculated using phyloseq (McMurdie and Holmes 2013) v.1.34.0 library in R. Differences in alpha diversity measures were assessed between pre and post FMT using the pairwise.wilcox.test base function in R with a Bonferroni correction for multiple comparisons. This function uses a paired, ranked, Mann-Whitney test to evaluate differences.

Principal coordinate analysis (PCoA) was performed to visualize the differences in relative abundance of pre and post FMT microbiota. Differences between pre and post FMT were evaluated by permutational multivariate analysis of variance (permANOVA) in the vegan package (Dixon 2003) v.2.5.7 of R.

## RESULTS

### Participants

We recruited 10 patients undergoing FMT to treat rCDI infections, of these 90% had at least 2 recurrences of rCDI before FMT. Of the 10 participants recruited for the study, 6 completed both visits, while 4 did not return for the 60-day follow-up visit after FMT. All patients had zero CDI recurrences and zero hospitalizations in the 60-day follow-up period post FMT. Additionally, FMT prevented further CDI recurrences for at least 2 years. However, though every patient was clinically cured with an absence of diarrhea and negative CDI test, of the 6 that completed both visits, two patients had persistent symptoms consistent with post infectious irritable bowel syndrome (IBS) as defined by Rome IV criteria (Drossman and Hasler 2016). The 6 participants that completed the second visit were female. The clinical characteristics are summarized in Table 1.

### FMT Increased Colonic Type 2 Cytokines

To evaluate changes in immune cell signaling in response to FMT, we measured 47 cell signaling proteins in colonic biopsy tissue lysates (Supplemental Table 1). IL-25 and IL-4 cytokines were of particular interest since they were related to eosinophil recruitment and a type 2 immune response, which we predicted *a priori* to be restored by FMT (Buonomo, Cowardin et al. 2016). IL1 has also been shown to be important in determining type-1 vs type-2 immune responses, so IL-1a and IL-1b were also selected *a priori* for analysis in the trial outcomes.

We found that protein concentrations of IL-1a, IL-1b, and IL-25 were significantly increased after FMT (linear mixed effect model, p<0.05) with a trend to increased IL-4 concentrations after FMT (p = 0.058) (Figure 1). Taking out the two patients that developed IBS-like symptoms, we found that the increase in IL-1a and IL-1b were no longer significant (p>0.1, p>0.05, respectively) and IL-4 increase was still not significant (p>0.1), while the increase in IL-25 was still significant (p<0.05), suggesting IBS-like symptoms may influence the concentrations of specific cytokines.

**Figure 1.**
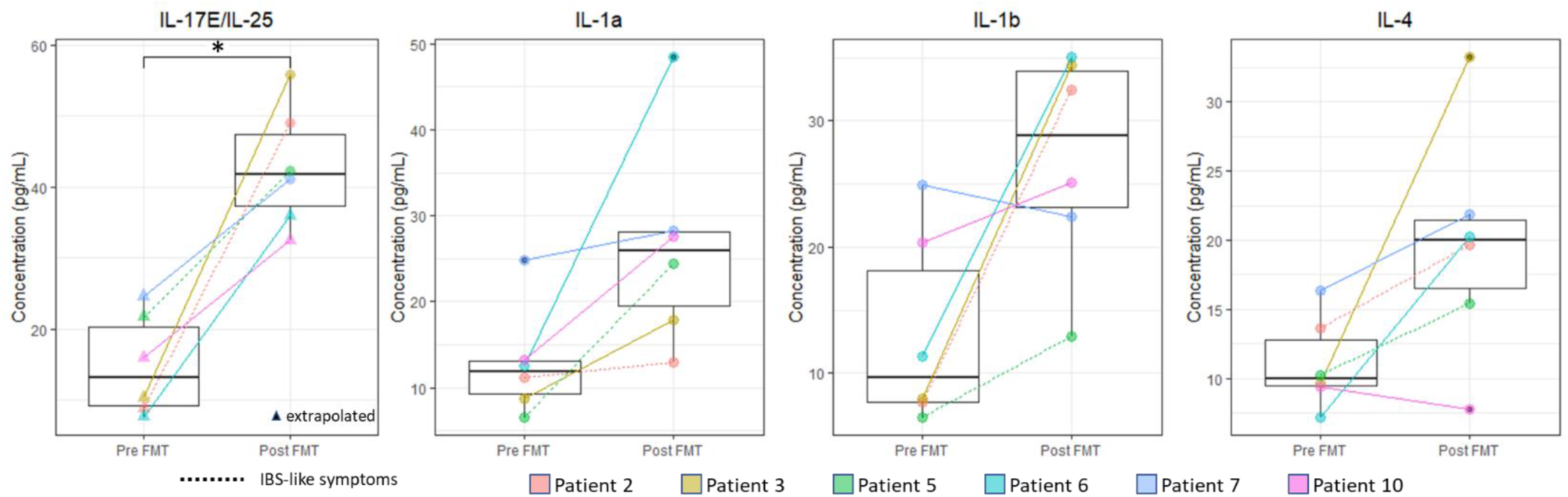
FMT increased Th2 cytokine levels in the colon. Levels of tissue cytokines were quantified by Luminex assay. The median value and IQR of tissue cytokines before and after FMT are shown. Linear mixed effect model with patient as a random variable was the statistical test used to calculate p-values. There were no significant differences in total protein concentration between pre and post FMT in the samples used for the Luminex assays (not shown). MFIs outside the standard curve range were extrapolated (triangles).

In the parallel transcriptomic analysis, while IL-25 transcripts could not be identified, FMT modestly increased the expression of *IL1a* (log_2_FC = 0.83) and *IL1b* (log_2_FC = 0.47) though these increases were not significant (Supplemental Figure S1A and B) and none of these cytokines were identified as differentially expressed genes. There was an increase in the transcript levels of the IL1 inhibitor IL1RA (encoded by *IL1RN*, log_2_FC = 0.83) following FMT (Supplemental Figure S1C). *IL1A* transcripts were negatively correlated with IL-1a protein levels (r<-0.6), while *IL1B* transcripts were positively correlated with IL-1b protein levels (r>0.6) (Supplemental Figure S1D). *IL1RN* transcripts were positively correlated with IL1RA protein concentration (r> 0.7) (Supplemental Figure S1D).

### Transcriptional Responses to FMT

To assess the transcriptional response in intestinal epithelial cells we compared gene expression in colonic biopsies obtained immediately prior to FMT to those obtained 60 days post FMT. PCA of the normalized gene counts from RNA-seq was done to investigate unsupervised groupings by visit, patient, or development of IBS-like symptoms post FMT (Figure 2A). Principal component 2 (PC2) separated the samples into pre vs post FMT while PC1 was related to patients that developed IBS-like symptoms post FMT-like symptoms. Negative loading unique to PC1 was driven by *OLFM4* (Figure 2B). *OLFM4* encodes olfactomedin-4, a selective marker of inflammation in the colonic epithelium (Liu and Rodgers 2016). The largest positive loading, *LYZ*, encodes for lysozyme which has been found previously to be expressed at lower levels in IBS patients relative to healthy controls (Aerssens, Camilleri et al. 2008). The top loadings uniquely driving PC2 were *ST6GAL2* (negative) and *HLA-DRB5* (positive). *ST6GAL2* encodes a sialyltransferase. Specifically in relation to the gastrointestinal system, sialic acid catabolism has been shown to be related to intestinal inflammation and microbial dysbiosis (Krzewinski‐Recchi, Julien et al. 2003, Huang, Chassard et al. 2015, Kim 2020) though the specific role of ST6GAL2 has not yet been established. *HLA-DRB5* encodes for an HLA class II protein which is specific to antigen presentation for immune cells and is related to ulcerative colitis (Silverberg, Cho et al. 2009). Three genes overlapped between PC1 and PC2 loadings: *MTRNR2L12, MTRNR2L8*, and *MTRNR2L2*, which encode isoforms of the mitochondrial peptide humanin, which has been shown to have cytoprotective effects (Bodzioch, Lapicka-Bodzioch et al. 2009).

**Figure 2.**
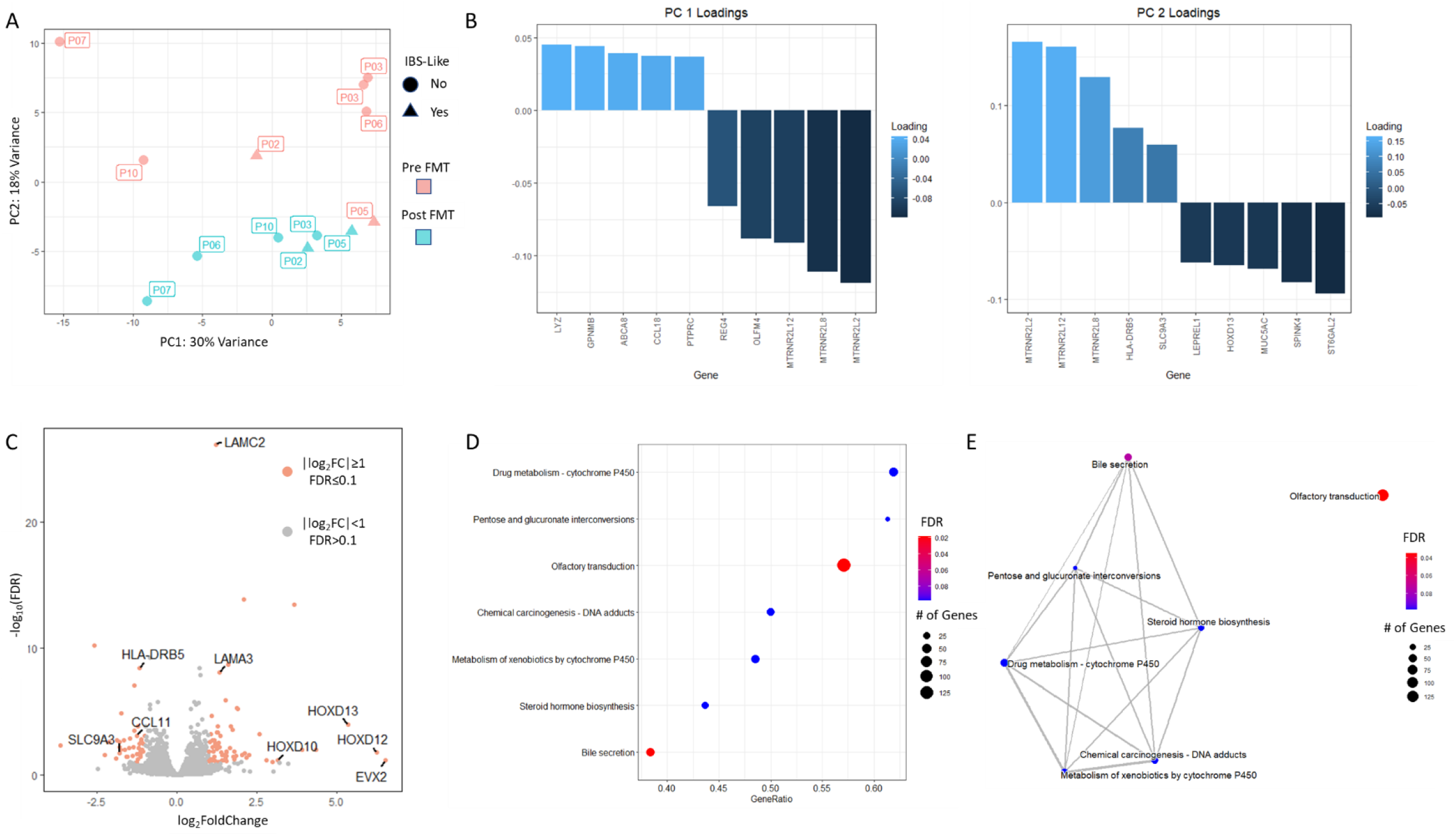
FMT driven variation in the colonic transcriptome. **A)** Principal component analysis of gene expression data identifies separation between pre (red) and post (green) FMT samples. Batch effects were minimal, as noted by how closely pre FMT patient 3 (P03) samples from 2 different batches grouped. PC1 captured some variance induced by the development of IBS-like symptoms post FMT in patients 2 (P02) and 5 (P05) clustered on the negative side of PC 1. PC 2 captured the variance related to FMT, where each patient consistently had a relatively higher PC2 value before FMT and a lower PC2 value after FMT. **B)** PC1 loadings were driven by *OLFM4* (positive) and LYZ (negative) while PC2 loadings were driven by *HLA-DRB5* (positive) and *ST6GAL2* (negative). **C)** Volcano plot of differentially expressed genes after FMT. Orange indicates genes that met the threshold of log_2_FC≥1, FDR≤0.1 (Wald test, Benjamini-Hochberg adjustment). Genes mentioned in the main manuscript are labeled here, which include homeobox genes, laminins, and some genes related to immune responses. **D)** Overrepresented pathways from pre to post FMT related to bile acids had overlapping genes. Using the FCs from DEGs from pre to post FMT, overrepresented pathways were found using KEGG gene lists (enrichment score test, Benjamini-Hochberg adjustment). **E)** Overlaps in pathways were mapped to find common themes.

We also performed hierarchical clustering of the 30 most variable genes before and after FMT (Figure S2). The dendrogram branched first between a group without any patients that developed IBS-like symptoms post FMT-like symptoms (right) and a group that included the two patients that developed IBS-like symptoms (left). For the no IBS-like symptom group, the second level of hierarchy was the split between pre and post FMT samples, before splitting by patient. The group that contained samples from patients that developed IBS-like symptoms were then split by patient before splitting into pre vs post FMT. The two patients that developed IBS-like symptoms post FMT did not cluster together suggesting patient-specific transcriptional responses vary even within patients with similar IBS-like symptoms.

### FMT induces transcriptional programs of tissue repair and suppresses inflammatory responses

We analyzed differentially expressed genes (DEGs) between pre and post FMT using a grouped model comparing all patients before and after FMT. Based on the unsupervised analyses of PCA (Figure 2A) and hierarchical clustering (Supplemental Figure 2) we incorporated patient ID into the model to account for patient-to-patient variability. A total of 103 DEGs were identified (log_2_FC≥1, FDR≤0.1) of which 62 were induced while 41 were repressed by FMT (Figure 2C). Some notable induced genes include homeobox genes (*EVX2, HOXD10, HOXD12, HOXD13*) and laminin genes (*LAMC2* and *LAMA3*). Homeobox genes have been shown to be related to development and differentiation of the intestinal epithelium (Beck 2002, de Santa Barbara, Van Den Brink et al. 2003), while laminins are components of the basal lamina which have been shown to be absent in ulcerative colitis (Teller and Beaulieu 2001). Some notable repressed genes include *HLA-DRB5, CCL11*, and *SLC9A3. HLA-DRB5* is a class II allele involved in antigen presentation to CD4 T cells and appeared in the above PCA analysis as being a positive loading to a component separating pre and post FMT. *CCL11*, also known as eotaxin-1, is pro-inflammatory and involved in eosinophil chemotaxis (Figure 2C) (Menzies-Gow, Ying et al. 2002). *SLC9A3* is related to bile acid elimination and diarhea (Hegyi, Maléth et al. 2018).

### FMT suppresses bile acid secretion and olfactory transduction pathways

To identify functional differences in gene expression due to FMT, the differentially expressed genes (DEGs) between pre and post FMT were analyzed for overrepresented pathways. The DEGs from the grouped model controlling for patient differences was used because this model was representative of significant gene expression differences common across all patients. Gene set enrichment analysis was analyzed in the KEGG human pathway database (Kanehisa and Goto 2000). A total of 7 pathways were significantly overrepresented (FDR < 0.1) among genes suppressed after FMT (Figure 2D). Functional overlaps in enriched pathways were identified for 6 of the 7 pathways found (Figure 2E). The overlapping pathways were (in order of decreasing gene set size): bile secretion, metabolism of xenobiotics by cytochrome P450, chemical carcinogenesis – DNA adducts, drug metabolism – cytochrome P450, steroid hormone biosynthesis, and pentose and glucuronate interconversions. Bile secretion and cytochromes P450 are related to oxidative stress, toxin elimination, and drug processing (Gonzalez and Gelboin 1994, Lorbek, Lewinska et al. 2012). Bile secretion can also be directly regulated by monooxygenases like cytochromes P450 (Lorbek, Lewinska et al. 2012). The repression of these pathways indicates a heightened transcriptional response against foreign invaders or the build-up of reactive oxygen species before FMT and a return to homeostasis after FMT. Of these overlapping pathways, the two most significant pathways were the bile secretion pathway (FDR = 0.074 enrichment score = -0.65) and the olfactory transduction pathway (FDR = 0.035, enrichment score = -0.74), which was not functionally linked to any of the other overrepresented pathways. Secondary bile acids have been shown to inhibit cell division of *C. difficile* (Kang, Myers et al. 2019). It has been previously shown that olfactory receptors are present all along the gastrointestinal tract and can be modulated by the intestinal microbiota (Priori, Colombo et al. 2015). The activation of odorant receptors has been linked to the inhibition of cell proliferation and apoptosis(Weber, Al-Refae et al. 2017).

### Peripheral Th17 subsets are decreased following FMT

To complement the whole tissue RNA-seq analyses, we used high dimensional flow cytometry to profile immune cell subsets in colonic biopsies and blood from before and after FMT. Lamina propria mononuclear cells (LPMCs) were isolated from colonic biopsies, and peripheral blood mononuclear cells (PBMCs) were isolated from venous blood samples. Overall, we isolated an average of 3291+/-2481 (mean +/-SD) viable CD45+ cells from the LPMCs from each patient, and an average of 255397 +/-80841 (mean +/-SD) viable CD45+ cells from the PBMCs. t-SNE plots were used to visualize cell populations in LPMC and PBMC samples and did not reveal any immune cell subsets unique to pre or post FMT (Supplemental Figure S2). Statistical analyses could not be used to compare cluster to cluster between pre and post FMT as specific subsets of interest appeared in various proportions overlapping one another and tSNE does not account for differences in the relative number of cells. Therefore, we used traditional gating to test for differences in subsets of interest. This revealed a significant decrease (p = 0.05, linear mixed effect model with patient as random variable) in the peripheral Th17 (CD3+CD19/CD20-CD4+RORyt+) after FMT (Figure 3L), a similar trend was noted in LPMCs but this did not reach statistical significance (Figure 3E). Despite this increase, there were no statistically significant differences in Th1:Th2, Th17:Treg ratios in LPMCs or PBMCs (not shown).

**Figure 3.**
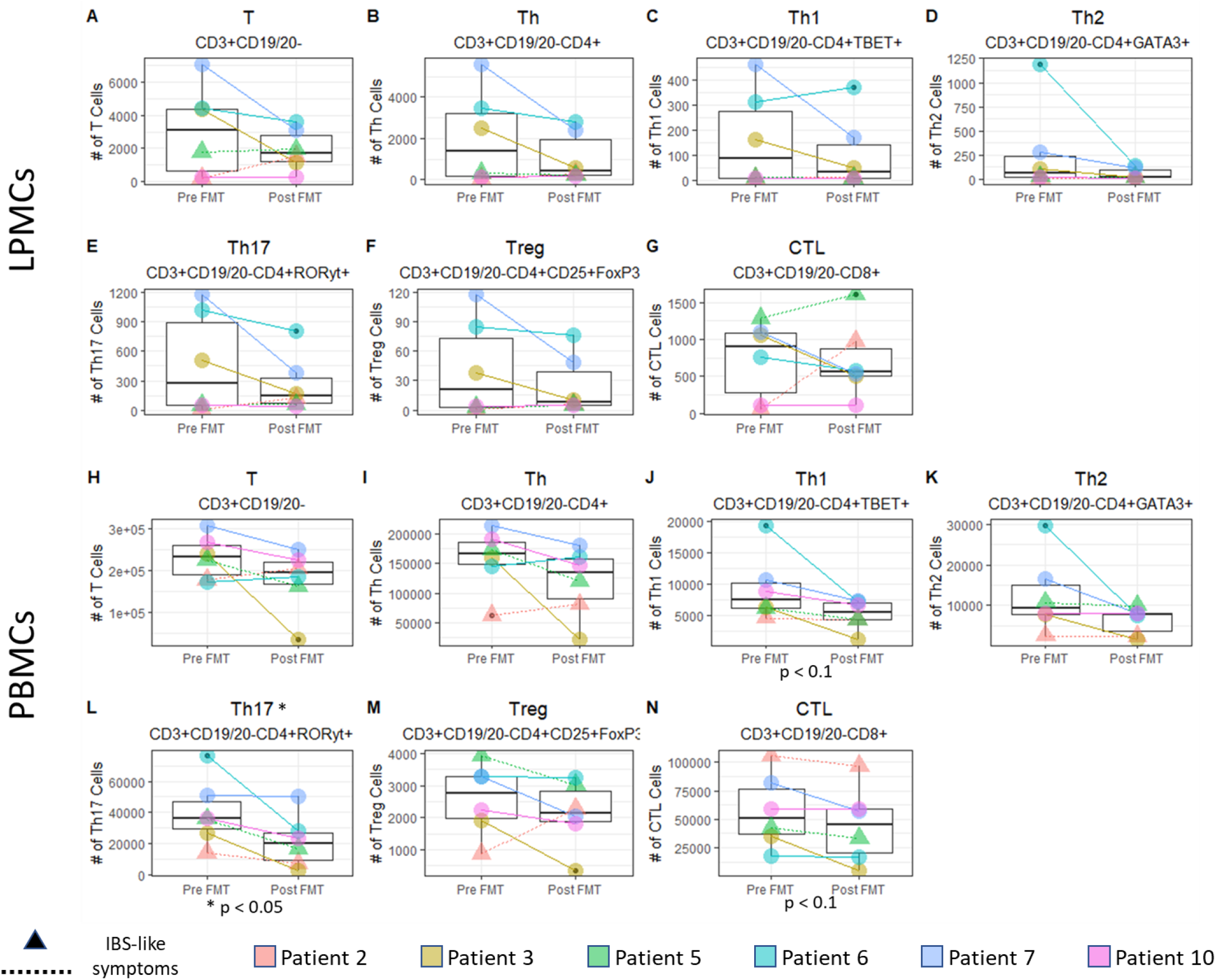
Peripheral Th17 cells were decreased after FMT (p < 0.05). A decrease in Th17 cells was observed for all patients except for patient 7, who had about the same number of Th17 cells pre and post FMT. The number of Th1 and cytotoxic T cells (CTL) were decreased post FMT, however these decreases were not statistically significant. There were no significant differences in the number of viable CD45+ cells between pre and post FMT nor were there significant differences in Th17 cells between patients that developed IBS-like symptoms post FMT and those that did not (p > 0.1). A linear mixed effect model with patient as a random variable was used to calculate p values.

### FMT increased microbial diversity

To evaluate the local changes in the microbial population by FMT, 16s rDNA sequencing was performed using DNA isolated from biopsy samples. The use of a biopsy allows us to investigate the mucosal microbial population in close association with the colonic epithelium, as opposed to the lumen of the gastrointestinal tract. We found that the alpha diversity measures Shannon and Simpson indices significantly increased after FMT (Mann-Whitney, p < 0.01) (Figure 4A and B), which agreed with other studies on diversity changes after FMT treatment for rCDI (Chang, Antonopoulos et al. 2008, Staley, Kaiser et al. 2018). For the beta diversity measures, clear groupings between pre and post FMT samples were evident in a PCoA plot (Figure 4C). The PCoA axis 1 explained 26.4% of the variance while axis 2 explained 16.9% and separated the patients by FMT status (permANOVA, alpha = 0.01). Patients that developed post IBS-like symptoms following FMT did not appear distinct in the PCoA analysis.

**Figure 4.**
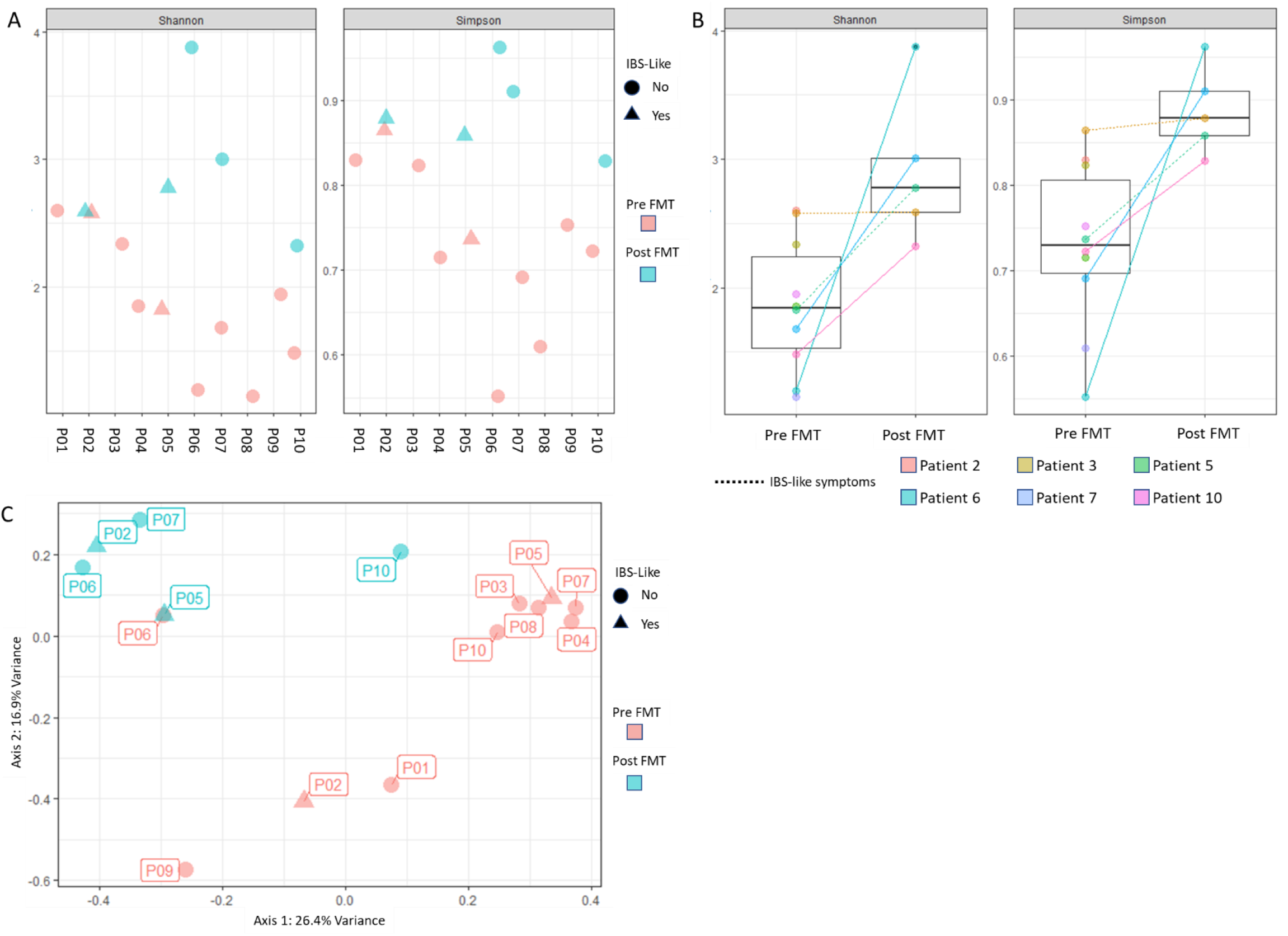
FMT increased alpha diversity of the epithelial-associated microbiome. **(a)**Increased Shannon and Simpson**(B)** Alpha Diversity measures were observed in 4 of the 5 patients analyzed (p < 0.01, Mann-Whitney). Patient 2 (P02) showed no change in diversity post FMT and developed IBS-like symptoms post FMT. **(C)** PCoA plot shows separation of relative abundance of microbiota populations before and after FMT. This separation was statistically significant (p < 0.01, permANOVA). One point that overlapped between pre and post FMT was from a pati1en1t (P5) that developed IBS-like symptoms post FMT.

The top 20 most abundant ASVs across all samples were grouped by taxonomic information and ranked. In the most abundant ASVs, the top families were *Enterobacteriaceae, Akkermansiaceae, Bacteroidaceae, Clostridiaceae, Acidaminococacear, Rikenellaceae, Fusobacteriaceae, Lachnospiraceae, Ruminococcaceae*, and *Campylobacteraceae*, in decreasing order of abundance. The top genera from the same list of ASVs were *Escherichia/Shigella, Akkermansia, Bacteroides, Clostridium_sensu_stricto_13, Phascolartobacterium, Alistepes, Lachnoclostridium, Clostridium_sensu_stricto_1, Agathobacter, Faecalibacterium, Fusicatenibacter, Campylobacter* and *Cetobacterium*, in decreasing order of abundance. Some notable taxa include the genera *Escherichia/Shigella, Akkermansia* and *Bacteroides*, which had different abundances between pre and post FMT. *Escherichia/Shigella* was found to be more abundant prior to FMT, which agrees with previous studies (Ling, Liu et al. 2014, Li, Gao et al. 2018). *Akkermansia* decreased after FMT while *Bacteroides* was increased after FMT. *Akkermansia muciniphila* plays a role in degrading mucins in the gut and is generally thought to have a beneficial effect on the gut to reduce inflammation (Schneeberger, Everard et al. 2015, Li, Lin et al. 2016, Zhao, Liu et al. 2017). *Bacteroides* has also been shown to have beneficial effects on the gut and has been shown to increase after FMT. *C. difficile* has also been shown to reduce *Bacteroides* growth (Khoruts, Staley et al. 2021). All patients were treated with vancomycin to treat *C. difficile* before FMT (Table 1) which may have had impacted the microbiota. A total of 15 ASVs were found to be differentially abundant between pre and post FMT (FDR < 0.05, Table 3). All differentially abundant ASVs were also present as one of the top 20 most abundance ASVs. The average transcriptional fold change of genes related to bile secretion was positively correlated (r>0.5) to differences in *Akkermansia* (Supplemental Figure S1E). *Akkermansia* abundance was negatively correlated (r <-0.7) with IL-25 protein concentration (Supplemental Figure S1F).

**Table 3.** Differentially abundant microbiota sorted by decreasing FDR (Wald test with Benjamini-Hochberg adjustment). “NA” indicates that the ASV for that microbe did not pertain to a specific species within that genus.

| log<sub>2</sub>FC | p value | FDR | Family | Genus | Species |
| --- | --- | --- | --- | --- | --- |
| -26.0 | 3.50E-16 | 1.61E-14 | *Akkermansiaceae* | *Akkermansia* | *muciniphila* |
| 23.2 | 1.21E-13 | 2.46E-12 | *Lachnospiraceae* | *Agathobacter* | NA |
| 23.1 | 1.60E-13 | 2.46E-12 | *Ruminococcaceae* | *Faecalibacterium* | NA |
| -23.1 | 5.39E-13 | 6.20E-12 | *Enterobacteriaceae* | *Klebsiella* | NA |
| -22.8 | 1.01E-12 | 9.33E-12 | *Clostridiaceae* | *Clostridium_sensu_stricto_1* | *paraputrificum* |
| 7.40 | 4.34E-03 | 0.0333 | *Rikenellaceae* | *Alistipes* | *putredinis* |
| 6.54 | 7.63E-03 | 0.0501 | *Bacteroidaceae* | *Bacteroides* | *vulgatus* |
| 7.95 | 0.0109 | 0.0570 | *Bacteroidaceae* | *Bacteroides* | *uniformis* |
| 7.80 | 0.0124 | 0.0570 | *Bacteroidaceae* | *Bacteroides* | *caccae* |
| 7.04 | 0.0113 | 0.0570 | *Lachnospiraceae* | *Anaerostipes* | *hadrus* |
| 7.25 | 0.0203 | 0.0778 | *Lachnospiraceae* | *Agathobacter* | NA |
| 7.33 | 0.0190 | 0.0778 | *Bacteroidaceae* | *Bacteroides* | *finegoldii* |
| 7.00 | 0.0251 | 0.0887 | *Bacteroidaceae* | *Bacteroides* | NA |
| -5.02 | 0.0309 | 0.0947 | *Clostridiaceae* | *Clostridium_sensu_stricto_13* | *subterminale* |
| 6.77 | 0.0301 | 0.0947 | *Rikenellaceae* | *Alistipes* | *shahii* |

## DISCUSSION

The most important finding of this study was that the cytokine IL-25 was increased after FMT. This supports a model where FMT acts to protect from rCDI by restoration of commensal bacteria signaling to the innate immune system via colonic epithelial IL-25. Previously we had demonstrated in the mouse model of CDI that the expression of IL-25 was rescued by FMT, and that IL-25 protected from CDI via type 2 immune responses in the gut (Buonomo, Cowardin et al. 2016). The finding that IL-25 is significantly increased in rCDI patients following successful FMT validates work in mice on the importance of this cytokine in the restoration of homeostasis post FMT. IL-25 has been shown to induce Th2-associated phenotypes like eosinophilic infiltration and increased mucus production in the gastrointestinal tract (Fort, Cheung et al. 2001). IL-25 has also been shown to increase eosinophil numbers as a protective response during *C. difficile* infection (Buonomo, Cowardin et al. 2016).

Additional cytokines that were increased post-FMT included IL-1b and IL-4. IL-1 been shown previously to influence the role of CD4 T cells (Santarlasci, Cosmi et al. 2013) and in particular, IL-1b has been shown to enhance Th2 differentiation (Caucheteux, Hu-Li et al. 2016). IL-1b has also been shown to be induced by *C. difficile* toxins in mice (Cowardin, Buonomo et al. 2016). A previous study found that IL-1b levels increased after vancomycin treatment (Ha, Kong et al. 2015), though this effect did not persist after vancomycin was discontinued. IL-4 has been shown to be important in Th2 responses, with IL-4 transcription being necessary for Th2 responses in mice (Kopf, Le Gros et al. 1993, Junttila 2018).

FMT treatment of rCDI patients also had significant effects on the epithelial transcriptional response, circulating immune cell population, cell-signaling proteins, and the mucosal microbiota. FMT induced the transcription of genes related to gut maintenance and integrity, including homeobox genes and laminins, and repressed the transcription of immune-related genes (*CCL11* and *HLA-DRB5*), detoxification (related to bile secretion and cytochromes P450 pathways), or apoptotic genes (related to olfactory transduction pathway), dampening the tissue inflammation.

Th17 cells in peripheral blood were decreased post-FMT. We have previously demonstrated in the mouse model of CDI that Th17 cells exacerbated CDI in part through the production of IL-17A and recruitment of neutrophils to the colon (Saleh, Frisbee et al. 2019). Other studies (Saleh, Frisbee et al. 2019, Cook, Rees et al. 2021, Cook, Rees et al. 2021) have also seen associations between Th17 cells and CDI, where Th17 cells were found to be significantly reduced in patients with rCDI versus new onset CDI, and that these Th17 cells were increased after FMT. This disagrees with our finding that FMT decreases the number of Th17 immune cells, though we only saw this decrease peripherally and not locally in the colonic tissue cells. It is important to note that this previous study focused on toxin B-specific T cells as opposed to total T cells. The differences between these previous findings and our study underscore that there may be distinct differences between the immune responses of toxin-specific immune cells and the overall state of the immune system during rCDI, or an immune system in CDI remission due to vancomycin treatment.

Two general themes emerge from our results. One is that after FMT, there is a transcriptional signature of dampened inflammatory immune responses in the colon. This is evidenced by the repression of key immune response-related chemokines, the suppression of pathways related to detoxification, and the decreased olfactory signal transduction which can be used to detect toxins or to inhibit apoptosis. This seems to be partially contradicted by the decrease in *Akkermansia muciniphila*, since this bacteria is generally inversely associated with inflammation (Earley, Lennon et al. 2019). This may be a result of vancomycin treatment prior to FMT. Studies have shown that vancomycin can increase the abundance of *Akkermansia muciniphila*, which can decrease *C. difficile* infection (Hansen, Krych et al. 2012, Ray and Aich 2019, Vakili, Fateh et al. 2020).

The second theme is the progression towards healing post FMT, as evidenced by the induction of homeobox and laminin genes related to cell proliferation as well as the increase in IL-25 tissue cytokines, which indicate a type-2 anti-inflammatory response. IL-4, another marker of a type-2 anti-inflammatory response was also found to be increased post FMT, though this was not significant. This increase in cytokine concentration did not match an increase in the transcriptional response of eosinophil-related processes though, so we cannot be sure if this increase is directly related to eosinophil recruitment. The increase in alpha diversity and *Bacteroides* are also a sign of healing post FMT, as previous studies have shown that an increased microbial diversity and increased abundance of *Bacteroides* is related to the success of FMT for treating *C. difficile* (Landy, Al‐Hassi et al. 2011, Deng, Yang et al. 2018).

One of the limitations of our study was we could not decouple the effects of CDI recovery from that of FMT only as it is not feasible to recruit healthy patients with no history of CDI for the same FMT procedures. Future studies may be supplemented by animal models in order to aid in differentiating between these conditions. However, our study still analyzes data directly from clinical patients and can analyze the effects of FMT treatment for rCDI in individual patients.

Another limitation of our study was the experimental challenges associated with using colonic biopsies as the source of samples. It is difficult to acquire large biomass, which can affect subsequent analyses, including LPMC isolation and DNA amplification (Bowcutt, Malter et al. 2015, Tang, Jin et al. 2020). For flow cytometric analyses of the infiltrating local immune cell population, the number of cells isolated and analyzed from the LPMCs derived from biopsies were low compared to the cells isolated from the blood. For one patient, the total number of viable CD45+ cells were below 500 for both pre and post FMT samples. To control for these differences, we analyzed immune cell phenotype differences using a mixed effect model that accounts for patient-to-patient differences. The microbiota analysis also suffered from low recovery of bacterial DNA relative to fecal samples, but an advantage of our analysis of epithelial associated species is a more accurate snapshot of the microbiota in close proximity to the epithelium in the specific region of the colon where FMT is expected to act (Tang, Jin et al. 2020).

In addition to the previous limitations, our study was also limited to the small number of patients that completed the study. Only 6 of the 10 originally enrolled patients completed the study by coming back 60 days post FMT. To account for this, when appropriate, unpaired patient analyses collected from pre FMT were still considered in the analyses. For example, in our 16 S rDNA analyses of microbiome differences between pre and post FMT, our beta diversity test on differences between pre and post FMT included diversity measures from patients that did not complete the study. However, the inclusion of pairwise analyses were able to account for patient-to-patient variability. This variability was larger in transcriptomic analyses compared to host microbiome compositional analyses. These differences emphasize that generalizations may need to take patient-to-patient differences into account.

## CONCLUSIONS

From our pairwise analyses, we found that rCDI patients had transcriptional evidence of dampened inflammatory responses and increased cell proliferation and healing after FMT. The exact relationships between transcriptomics, immune cell population, and microbiota may be different from person to person due to the complex nature of these interacting components of intestinal health. Our analyses show that population trends were evident in microbiota compositional analyses but incorporating relative differences by patient increased the sensitivity of the transcriptional analysis due to higher inter-patient variability. The finding that FMT increased the concentration of IL-25 indicate that anti-inflammatory eosinophil recruitment may be part of the mechanism behind FMT treatment of rCDI. The role of IL-25 in restoring mucosal homeostasis merits further investigation as a potential adjuvant to FMT for treatment of rCDI.

## Data Availability

The authors confirm that the data supporting the findings of this study are available within the article and its supplementary materials. Any other data are available from the corresponding author, CM, upon reasonable request.

## ACKNOWLEDGEMENTS

Research reported in this publication was supported by National Institute of Allergy and Infectious Diseases of the National Institutes of Health under award numbers R01 AI124214 and R01 AI152477 to WP. We also thank the UVA Genomics Core Facility in the Department of Biology for sequencing and library preparation for 16 S rDNA microbiota analyses (RRID:SCR_012197), the UVA SOM Flow Cytometry Core for aiding with flow cytometry and Luminex assay and the Bioinformatics Core for their help with 16 S rDNA taxonomic analyses (University of Virginia Strategic Investment Fund #162 TransUniversity Microbiome Initiative).

