## Supplementary Figures and Tables for "Fecal microbiota transplantation increases colonic IL-25 and dampens tissue inflammation in patients with recurrent *Clostridioides difficile*"

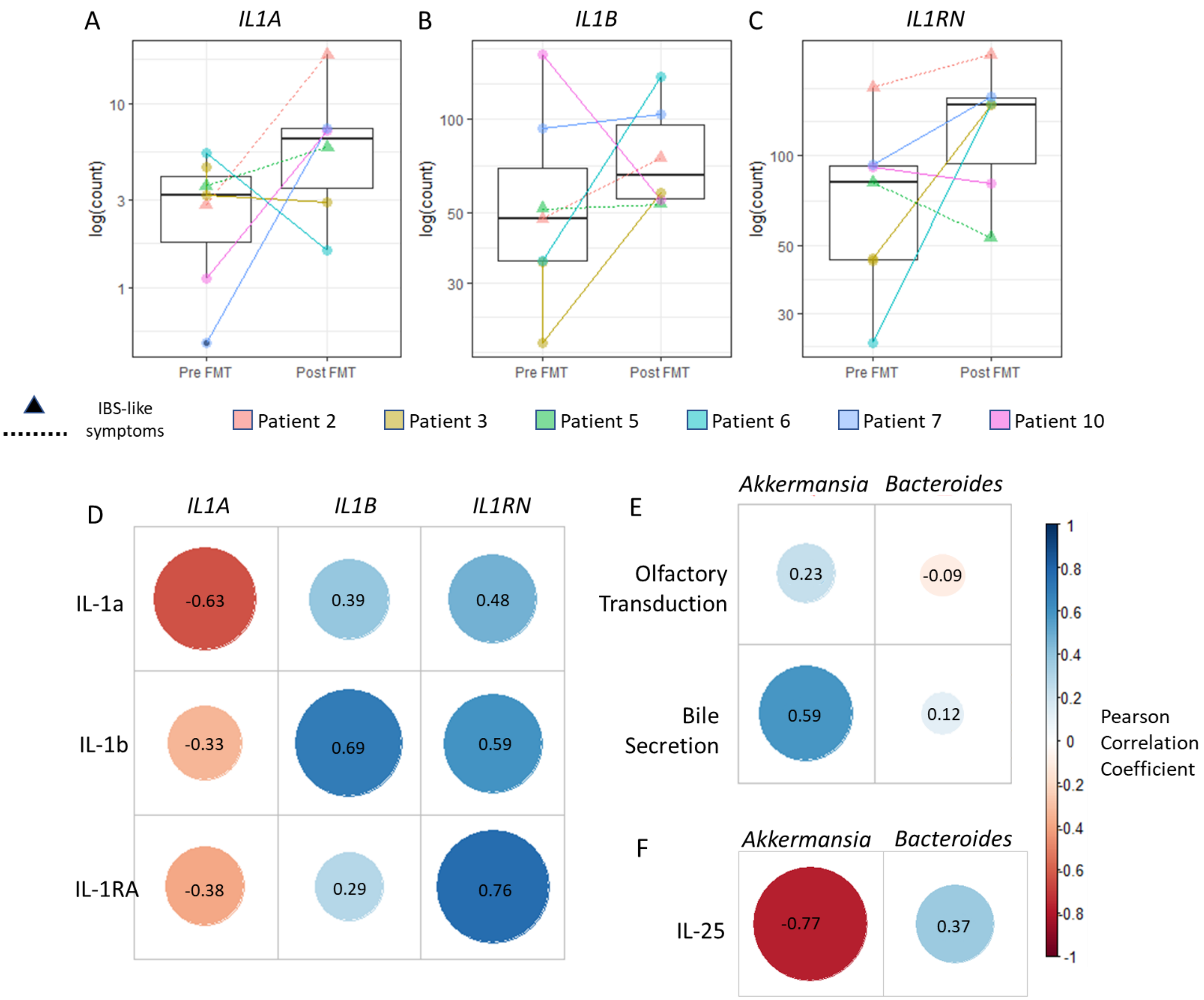


**Supplemental Figure S1. Correlations between different analyses show some relationships between gene transcription, protein concentration, and epithelial microbial abundance.** There was moderate induction of *IL1A* (A), *IL1B* (B), and *IL1RN* (C) transcription from pre to post FMT and these increases were not significant (p> 0.1, Wald test). For all three genes, the transcriptional response to FMT was variable from patient to patient. For the transcription of *IL1A*, some patients exhibited little change (patient 3) or a decrease from pre to post FMT (patient 6). For the transcription of *IL1B*, patient 10 exhibited a decrease in transcription, and there was little change in transcription for patients 5 and 7. For the transcription of *IL1RN*, patients 5 and 10 showed decreases in transcription. *IL1A* gene transcription had a negative correlation with IL-1a protein concentration, but for genes *IL1B* and IL1RN, there were positive correlations with their respective proteins. *Akkermansia* genus had a positive correlation with bile secretion pathway genes (E) and a negative relationship with IL-25 protein concentration (F).

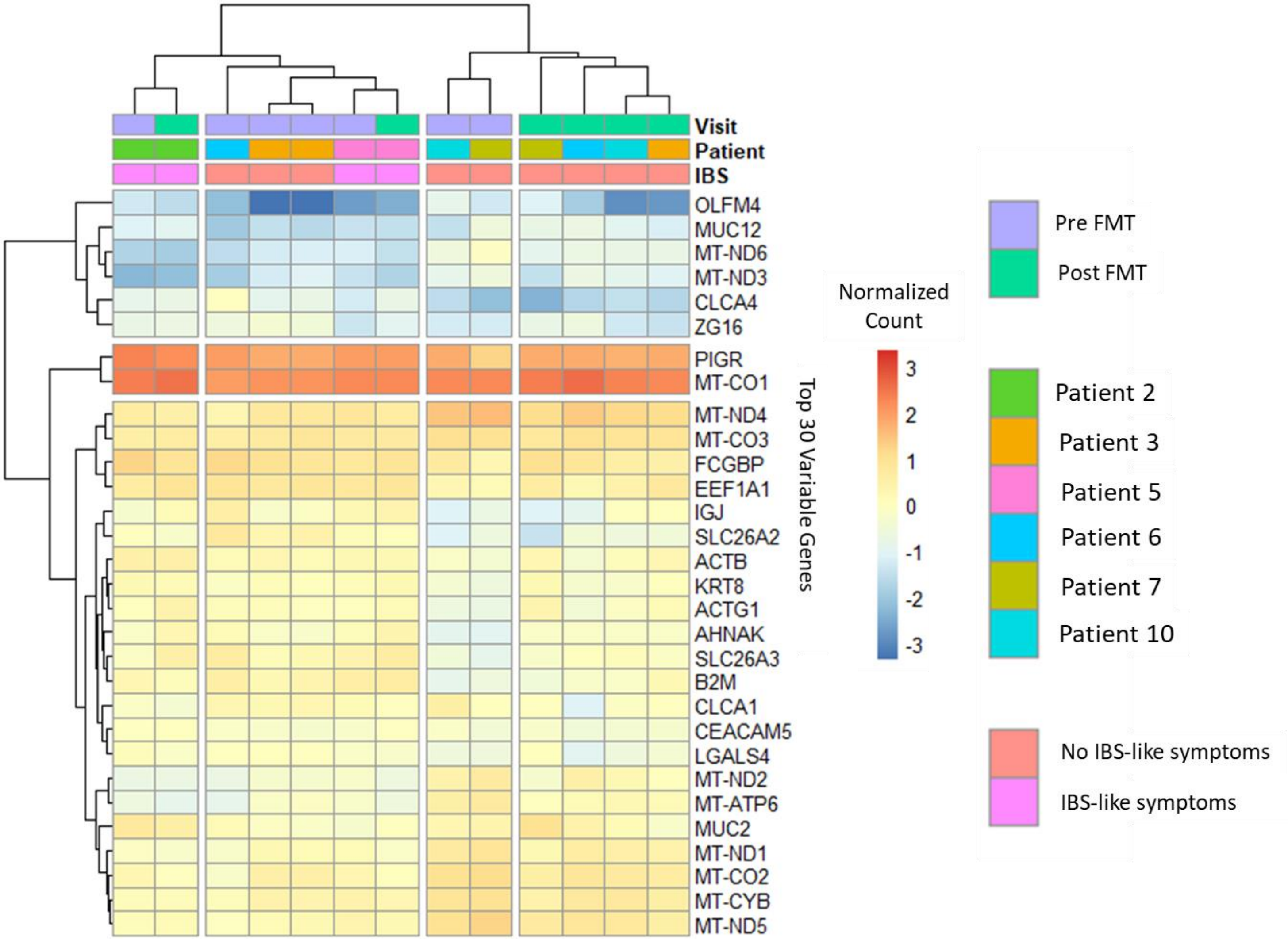


**Supplemental Figure S2. Hierarchical clustering of the top 30 variable genes by standard deviation of RNA-seq counts show some separation between patients that developed IBS-like symptoms post FMT and those that did not.** Colored boxes near the top of the plot indicate the visit (purple for pre FMT and green for post FMT), patient ID, and whether the patient developed IBS-like symptoms post FMT (orange for no IBS-like symptoms, and pink for developing IBS-like symptoms), while below that is a heatmap of the normalized counts scaled around zero, where lower counts are blue and higher counts are red. In the first level of hierarchical clustering, the samples were separated into a group consisting of 5 samples from patients that did not develop IBS-like symptoms post FMT and another that included all samples from patients that did develop IBS-like symptoms post FMT.

LPMC pre FMT

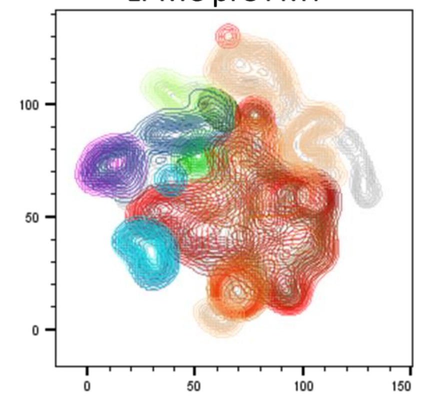


LPMC post FMT

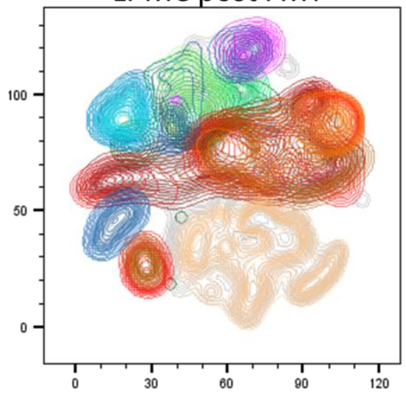


| | Subset Name | Count |
|---|---|---|
| | CD22+ B Cell (CD3-CD19_CD20+CD22+) | 926 |
| | B Cell (CD3-CD19_CD20+) | 2806 |
| | Cytotoxic T Cell (CD3+CD19_CD20-CD4-CD8+) | 4354 |
| | Treg (CD3+CD19_CD20-CD4+CD8-FOXP3+CD25+) | 247 |
| | Th17 (CD3+CD19_CD20-CD4+CD8-RORyt+) | 2803 |
| | Th2 (CD3+CD19_CD20-CD4+CD8-GATA3+) | 615 |
| | Th1 (CD3+CD19_CD20-CD4+CD8-TBET+) | 964 |
| | M2: resident macrophage (CD64+CD11c-CD163+CD11c-CD163hi) | 0 |
| | M1: inflammatory macrophage (CD64+CD14+CD11c+CD163lo) | 376 |
| | ILC3 (CD3-CD19_CD20-CD16-CD14-RORyt+GATA3-) | 628 |
| | ILC2 (CD3-CD19_CD20-CD16-CD14-RORyt-GATA3+) | 106 |
| | ILC1 (CD3-CD19_CD20-CD16-CD14-RORyt-GATA3-TBET+) | 25.0 |
| | Ungated | 24010 |

| | Subset Name | Count |
|---|---|---|
| | CD22+ B Cell (CD3-CD19_CD20+CD22+) | 575 |
| | B Cell (CD3-CD19_CD20+) | 1760 |
| | Cytotoxic T Cell (CD3+CD19_CD20-CD4-CD8+) | 4290 |
| | Treg (CD3+CD19_CD20-CD4+CD8-FOXP3+CD25+) | 150 |
| | Th17 (CD3+CD19_CD20-CD4+CD8-RORyt+) | 1574 |
| | Th2 (CD3+CD19_CD20-CD4+CD8-GATA3+) | 308 |
| | Th1 (CD3+CD19_CD20-CD4+CD8-TBET+) | 612 |
| | M2: resident macrophage (CD64+CD11c-CD163+CD11c-CD163hi)) | 1.00 |
| | M1: inflammatory macrophage (CD64+CD14+CD11c+CD163lo) | 211 |
| | ILC3 (CD3-CD19_CD20-CD16-CD14-RORyt+GATA3-) | 480 |
| | ILC2 (CD3-CD19_CD20-CD16-CD14-RORyt-GATA3+) | 77.0 |
| | ILC1 (CD3-CD19_CD20-CD16-CD14-RORyt-GATA3-TBET+) | 32.0 |
| | Ungated | 15642 |

PBMC pre FMT

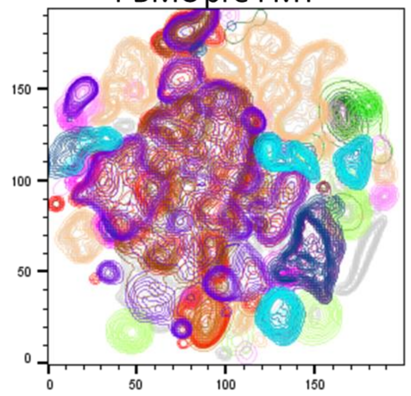


PBMC post FMT

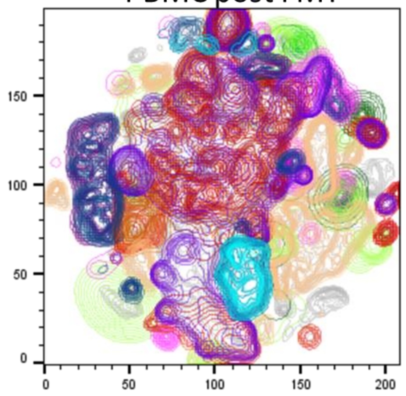


| | Subset Name | Count |
|---|---|---|
| | CD22+ B Cell (CD3-CD19_CD20+CD22+) | 92288 |
| | B Cell (CD3-CD19_CD20+) | 208734 |
| | Th1 (CD3+CD19_CD20-CD4+CD8-TBET+) | 55979 |
| | Cytotoxic T Cell (CD3+CD19_CD20-CD4-CD8+) | 341875 |
| | Treg (CD3+CD19_CD20-CD4+CD8-FOXP3+CD25+) | 15534 |
| | Th17 (CD3+CD19_CD20-CD4+CD8-RORyt+) | 240158 |
| | Th2 (CD3+CD19_CD20-CD4+CD8-GATA3+) | 75090 |
| | Non-Classical Monocytes (CD14-CD16+) | 15832 |
| | Intermediate Monocytes (CD14+ CD16+) | 18103 |
| | Classical Monocytes (CD14+ CD16-) | 102814 |
| | ILC3 (CD3-CD19_CD20-CD16-CD14-RORyt+GATA3-) | 1520 |
| | ILC2 (CD3-CD19_CD20-CD16-CD14-RORyt-GATA3+) | 2980 |
| | ILC1 (CD3-CD19_CD20-CD16-CD14-RORyt-GATA3-TBET+ | 723 |
| | Ungated | 1.71E6 |

| | Subset Name | Count |
|---|---|---|
| | CD22+ B Cell (CD3-CD19_CD20+CD22+) | 64593 |
| | B Cell (CD3-CD19_CD20+) | 188507 |
| | Th1 (CD3+CD19_CD20-CD4+CD8-TBET+) | 31445 |
| | Cytotoxic T Cell (CD3+CD19_CD20-CD4-CD8+) | 267799 |
| | Treg (CD3+CD19_CD20-CD4+CD8-FOXP3+CD25+) | 12767 |
| | Th17 (CD3+CD19_CD20-CD4+CD8-RORyt+) | 129466 |
| | Th2 (CD3+CD19_CD20-CD4+CD8-GATA3+) | 37143 |
| | Non-Classical Monocytes (CD14-CD16+) | 15185 |
| | Intermediate Monocytes (CD14+ CD16+) | 25955 |
| | Classical Monocytes (CD14+ CD16-) | 134021 |
| | ILC3 (CD3-CD19_CD20-CD16-CD14-RORyt+GATA3-) | 573 |
| | ILC2 (CD3-CD19_CD20-CD16-CD14-RORyt-GATA3+) | 1564 |
| | ILC1 (CD3-CD19_CD20-CD16-CD14-RORyt-GATA3-TBET+ | 414 |
| | Ungated | 1.36E6 |

**Supplemental Figure S2. tSNE groupings did not show any immune cell populations unique to pre or post FMT in both LPMCs and PBMCs.** The analyses were done separately between pre and post FMT in order to highlight any immune cell populations that were unique to pre or post FMT, here visualized as different colors. A contour map was used to show groupings within each immune cell subset. The color schemes and sizes of groupings were similar from pre to post FMT, showing no obvious differences in immune cell populations between pre and post FMT.

**Table S1. Pre vs Post FMT protein concentrations from Luminex assay. Linear mixed effect model with patient as random variable was used to calculate significance and a Benjamini-Hochberg method was used to adjust for multiple comparisons.**

| | | Pre FMT | | Post FMT | | | |
|---|---|---|---|---|---|---|---|
| Type | Analyte | Median (pg/mL) | IQR (pg/mL) | Median (pg/mL) | IQR (pg/mL) | *p* value | FDR |
| Chemokine | MCP-1 (CCL2) | 110.0 | 32.5 | 147.6 | 116.7 | 0.353 | 0.405 |
| | MIP-1α (CCL3) | 17.7 | 18.6 | 31.1 | 11.3 | 0.235 | 0.307 |
| | MIP-1β (CCL4) | 39.6 | 9.3 | 39.9 | 20.6 | 0.824 | 0.824 |
| | MCP-3 (CCL7) | 13.9 | 4.4 | 35.7 | 11.0 | 0.00950 | 0.102 |
| | EOTAXIN-1 (CCL11) | 25.3 | 15.2 | 27.0 | 7.9 | 0.455 | 0.497 |
| | MDC (CCL22) | 28.6 | 14.0 | 24.0 | 21.4 | 0.710 | 0.726 |
| | GROα (CXCL1) | 86.5 | 66.2 | 267.6 | 132.1 | 0.136 | 0.208 |
| | IL-8 (CXCL8) | 23.1 | 24.9 | 36.0 | 25.4 | 0.265 | 0.328 |
| | MIG (CXCL9) | 23799.5 | 17844.5 | 4073.0 | 8637.3 | 0.0183 | 0.102 |
| | IP-10 (CXCL10) | 379.0 | 432.9 | 62.9 | 95.6 | 0.147 | 0.209 |
| | Fractalkine (CX3CL1) | 72.9 | 11.8 | 169.4 | 56.0 | 0.0232 | 0.102 |
| Cytokine | Flt-3 ligand | 11.4 | 4.1 | 12.4 | 8.1 | 0.347 | 0.405 |
| | IFNα2 | 33.1 | 17.1 | 73.6 | 28.8 | 0.0161 | 0.102 |
| | IFNγ | 45.9 | 18.8 | 155.7 | 105.7 | 0.0347 | 0.102 |
| | **IL-1α** | 11.8 | 3.8 | 26.0 | 8.6 | 0.0424 | 0.102 |
| | **IL-1β** | 9.7 | 10.3 | 28.8 | 10.8 | 0.0315 | 0.102 |
| | IL-1RA | 1080.5 | 969.4 | 3475.5 | 1246.3 | 0.0218 | 0.102 |
| | IL-2 | 0.2 | 0.1 | 0.9 | 0.6 | 0.151 | 0.209 |
| | IL-3 | 2.1 | 13.3 | 37.5 | 4.5 | 0.0044 | 0.0689 |
| | **IL-4** | 10.0 | 3.3 | 20.0 | 5.0 | 0.0583 | 0.125 |
| | IL-5 | 0.8 | 0.2 | 2.2 | 1.1 | 0.0307 | 0.102 |
| | IL-6 | 1.6 | 0.7 | 2.9 | 1.6 | 0.147 | 0.209 |
| | IL-7 | 3.1 | 11.2 | 11.8 | 18.7 | 0.277 | 0.333 |
| | IL-9 | 30.1 | 7.9 | 91.7 | 30.6 | 0.0040 | 0.069 |
| | IL-10 | 149.2 | 87.9 | 441.4 | 242.1 | 0.0194 | 0.102 |
| | IL-12p40 | 15.4 | 19.4 | 21.1 | 9.1 | 0.642 | 0.670 |
| | IL-12p70 | 1.7 | 0.2 | 4.5 | 3.2 | 0.0214 | 0.102 |
| | IL-13 | 11.7 | 3.7 | 35.5 | 20.5 | 0.0257 | 0.102 |
| | IL-15 | 94.3 | 10.1 | 103.8 | 59.2 | 0.1077 | 0.181 |
| | IL-17A | 6.4 | 3.6 | 15.1 | 12.6 | 0.0400 | 0.102 |
| | **IL-17E (IL-25)** | 13.1 | 11.1 | 41.8 | 10.1 | 0.0016 | 0.069 |
| | IL-17F | 2.4 | 1.9 | 9.0 | 6.2 | 0.0324 | 0.102 |
| | IL-18 | 18.6 | 14.7 | 11.1 | 2.2 | 0.245 | 0.311 |
| | IL-22 | 142.1 | 188.0 | 317.9 | 80.4 | 0.0711 | 0.135 |
| | IL-27 | 849.1 | 1083.4 | 1514.0 | 430.5 | 0.635 | 0.670 |
| | TGF-α | 13.8 | 9.0 | 23.0 | 14.2 | 0.0746 | 0.135 |

| | | | | | | | |
|---|---|---|---|---|---|---|---|
| | TNF-α | 17.6 | 8.7 | 29.1 | 24.4 | 0.374 | 0.419 |
| | TNF-β | 4.1 | 1.4 | 15.0 | 11.0 | 0.0494 | 0.111 |
| | sCD40L | 4105.5 | 1472.5 | 6123.0 | 3395.8 | 0.0742 | 0.135 |
| Growth factor | EGF | 25.0 | 3.9 | 51.1 | 20.3 | 0.0344 | 0.102 |
| | FGF-2 | 9885.5 | 1934.5 | 13651.0 | 3927.5 | 0.0435 | 0.102 |
| | G-CSF | 76.0 | 92.9 | 157.1 | 343.3 | 0.137 | 0.208 |
| | GM-CSF | 0.0 | 0.2 | 1.6 | 1.6 | 0.0891 | 0.155 |
| | M-CSF | 476.8 | 80.1 | 683.8 | 926.6 | 0.120 | 0.194 |
| | PDGF-AA | 246.5 | 94.5 | 439.2 | 195.2 | 0.0652 | 0.133 |
| | PDGF-AB/BB | 0.0 | 0.0 | 123.0 | 167.5 | 0.0376 | 0.102 |
| | VEGF | 126.4 | 103.9 | 206.8 | 189.0 | 0.164 | 0.220 |

**Table S2. Flow Cytometry Panel**

| | Marker | Fluorophore |
|---|---|---|
| Surface Stains | CCR7 | BUV395 |
| | CD103 | BUV 563 |
| | CD83 | BUV 737 |
| | CD22 | BUV805 |
| | CD14 | Pacific Blue |
| | CD127 | eFluor506 |
| | CD11b | BV510 |
| | CD45RA | BV570 |
| | CD163 | BV605 |
| | CD4 | BV650 |
| | CD8 | BV750 |
| | HLA-DR | BV785 |
| | CD45 | QDOT 800 |
| | CD11c | BB515 |
| | CD3 | Spark 550 |
| | CD117 | PerCP-Cy5.5 |
| | gdTCR | PerCP-ef710 |
| | CD25 | PE-Dazzle 594 |
| | CD294 | PE-Cy5 |
| | CD16 | PE-Cy7 |
| | CD19/20 | APC-Cy5.5 |
| | CD56 | AF700 |
| | CD64 | APC-fire 750 |

| | Marker | Fluorophore |
|---|---|---|
| Intracellular Stains | FoxP3 | BV421 |
| | Tbet | BV711 |
| | Ki67 | AF532 |
| | Gata3 | PE |
| | RORyt | AF647 |
